# Readmissions in Patients with Cerebral Cavernous Malformations (CCMs): A National Readmission Database (NRD) Study

**DOI:** 10.1101/2021.09.18.21263780

**Authors:** Akhil Padarti, Amod Amritphale, Javed Khader Eliyas, Daniele Rigamonti, Jun Zhang

## Abstract

**BACKGROUND:** Cerebral cavernous malformations (CCMs) are microvascular CNS lesions prone to hemorrhage leading to neurological sequela such as stroke and seizure. A subset of CCM patients have aggressive disease leading to multiple bleeding events, likely resulting multiple hospitalizations. Hospital admission rates are an important metric that has direct financial impact on hospitals and an indicator of overall disease burden. Furthermore, analysis of hospital readmissions can lead to early identification of high-risk patients and provides insight into the pathogenesis of CCM lesions. The purpose of this study is to identify high risk CCM patients with increased all cause readmission and comorbidities associated with increased readmissions.

**METHODS:** All US hospital admissions due to CCMs were searched using the 2017 National Readmission Database (NRD). Patients with readmissions within 30 days of discharge from index hospitalization were identified and analyzed, relative to the remaining population.

**RESULTS:** Among all patients hospitalized for CCM, 14.9% (13.7-16.2%) required all cause readmission within 30 days. Multivariate logistical regression analysis showed that substance abuse (p=0.003), diabetes (p=0.018), gastrointestinal bleed (p=0.002), renal failure (p=0.027), and coronary artery disease (p=0.010) were predictive of all cause readmissions, while age group 65-74 (p=0.042), private insurance (p<0.001), and treatment at a metropolitan teaching institution (p=0.039) were protective. Approximately half of all readmissions are caused by neurological (33.9%) and infectious (14.6%) etiologies. The 30-day lesion bleeding rate after index hospitalization is 0.8% (0.5-1.2%).

**CONCLUSIONS:** All identified comorbidities associated with increased risks of readmission contribute to vascular stress, suggesting its role in lesion pathogenesis. This is the first and only study to analyze readmission metrics for CCMs in order to identify high risk patient factors to date.

## Introduction

Cerebral cavernous malformations (CCMs) are microvascular lesions predominantly found in the central nervous system, composed of dilated venous sinusoids with a singular layer of endothelium and minimal intervening brain parenchyma. CCMs are the second most common cerebral vascular abnormality following developmental venous anomaly (DVA) ^1^. It is estimated that CCM lesions are present in 1 in 200-600 in the general population with higher prevalence in Hispanic population (1 in 70) ^2–4^. These lesions are predisposed to hemorrhagic strokes and seizures ^5^. Patients exhibit a wide range of disease severity depending on the underlying gene mutation, lesion burden, location, and frequency of hemorrhage. A substantial number of patients with CCMs are asymptomatic (20-70%) ^6–8^.

However symptomatic patients can have devastating neurological deficits often due to CCM lesion bleeding. If the frequency of lesion bleeding can be reduced, then the quality of life for CCM patients could be drastically improved. It has been shown that patients with one bleeding event have an increased risk (>10%) for a recurrent bleeding event ^9, 10^. Therefore, one subset of CCM patients have asymptomatic lesions while another subset have aggressive lesions with multiple hemorrhages. Identifying common factors unique to patients with aggressive disease will not only help clinicians distinguish high-risk patients, but also the shared characteristics may provide insight into CCM lesion pathogenesis.

Readmission rates are important hospital metrics used to gauge quality of healthcare received and have direct financial implications ^11^. CCM patients with aggressive phenotypes are more likely to incur admissions and readmissions to hospitals. Analyzing readmission rates will help identify CCM patients with high lesion burden in efforts that may lead to decreasing future hospitalizations. To date, this is the only study on CCM readmission rates to identify high risk CCM patients. The purpose of this study is to identify high risk CCM patients with increased all cause readmission and associated comorbidities.

## Materials and methods

### National Readmission Database

This study examined CCM readmissions using hospital admissions data from the National Readmission Database (NRD), Healthcare Cost and Utilization Project (HCUP), Agency for Healthcare Research and Quality, for 2017 ^12^. This is largest publicly available national representative sample of all admissions and discharges from U.S. nonfederal hospitals provided by the Agency of Healthcare Research and Quality. It contains data from 28 geographically dispersed states across the U.S (www.hcup-us.ahrq.gov/hcupdatapartners.jsp). It has about 18 million discharges for the year 2017. This database contains hospitalizations from 60% of the U.S. population. A unique de-identified linkage number is assigned to each patient which tracks readmissions within the same state.

### Patient Determination

CCM was defined with the diagnosis codes from CCSR/ICD-10 (supplementary table 1). Index hospitalization is any patient non-electively admitted to the hospital with a primary admission diagnosis of CCMs. The primary outcome variable was the first non-elective readmission within 30 days after discharge from index hospitalization. Only the first of these readmissions was included in the analysis. Exclusion criteria include any admission in December 2017 as they were not followed up for at least 30 days (no access to 2018 data), patients that died during the initial hospitalization, or had missing data for discharge. The study used publicly available deidentified database and was approved for exempt status by the University of South Alabama Institutional Review Board (IRB). Our study followed the methodology as previously utilized in other studies ^13, 14^.

### Clinical Variables

Pertinent patient demographics were obtained by NRD variables: age, sex, weekend admission, insurance status, and quartile of household income (Table 1). The quartile of household income was determined by ZIP code demographic data. Cost analysis of the repeat admission was performed by analyzing data on index admission length of stay (LOS) (days), index admission cost (U.S. dollar), and discharge destination (Table 1). Hospital charges were multiplied with the Agency for Healthcare Research and Quality’s all-payer cost-to-charge ratios to determine costs. The burden of the CCM lesions was measured by risk of mortality and loss of function subclasses in the index hospitalization (Table 1). Baseline patient comorbidities during the index hospitalization were determined by CCSR/ICD-10 codes shown in supplementary table 1. The first diagnosis CCSR/ICD-10 code was determined to be cause of the readmission (Table 4). All CCSR/ICD-10 codes used in this study are presented in the supplemental section.

**Table 1:** CCM Patient Characteristics during Index Hospitalization for All Cause Readmission. Baseline patient characteristics from NRD are shown i.e. age, gender, weekend admission, true cost of hospitalization, length of stay, insurance status, quartile of median household income, APRDRG: likelihood of dying, APRDRG: loss of function, hospital bed size, hospital teaching status, discharge location, control/ownership of hospital, and hospital designation. Age, true cost of hospitalization, and length of stay is presented as median and interquartile range (IQR), while all other discrete characteristics are presented as percentages. Characteristics of patients without all cause 30-day readmission (first column), with all cause 30-day readmission (second column), and total patient population (third column) are shown. The p-value for the characteristic between the two groups is also shown (fourth column). All significant p-values are bolded. The cutoffs of quartile of median household income, APRDRG: likelihood of dying, APRDRG: loss of function, and hospital bed size were defined as per HCUP. All data shown is weighted values in order to maintain patient privacy. Abbreviations: IQR-Interquartile range, LOS-length of stay, APRDRG-all patient refined diagnosis related groups. *p<0.05, **p<0.01, ***p<0.001.

|  | No 30d readmission N (%) | 30d readmission N (%) | Overall N (%) | P Value |
| --- | --- | --- | --- | --- |
| **Age(in years), median [IQR]** | 58 (42-71) | 65 (49-77) | 58 (42-72) | 0.248 |
| **Women** | 1624 (56.4%) | 261 (51.8%) | 1885 (55.7%) | 0.053 |
| **Weekend admission** | 579 (20.1%) | 123 (24.4%) | 702 (20.8%) | **0.029\*** |
| **Cost of hospitalization in US\$, median [IQR]** | 9200 (5500-18700) | 10100 (6000-21500) | 9300 (5600-19200) | **<0.001\*\*\*** |
| **LOS, median [IQR]** | 3 (2-5) | 4 (2-7) | 3 (2-5) | **<0.001\*\*\*** |
| **Insurance status** |  |  |  | **<0.001\*\*\*** |
| Medicare | 1191 (41.4%) | 286 (56.9%) | 1477 (43.7%) |  |
| Medicaid | 395 (13.7%) | 82 (16.3%) | 477 (14.1%) |  |
| Private | 1071 (37.3%) | 99 (19.7%) | 1170 (34.6%) |  |
| Self-pay | 107 (3.7%) | 19 (3.8%) | 126 (3.7%) |  |
| No charge | 12 (0.4%) | - | 12 (0.4%) |  |
| Other | 98 (3.4%) | 17 (3.4%) | 115 (3.4%) |  |
| **Quartile of median household income** |  |  |  | 0.167 |
| 0-25th | 672 (23.6%) | 134 (27.3%) | 806 (24.1%) |  |
| 26th-50th | 840 (29.5%) | 134 (27.3%) | 974 (29.2%) |  |
| 51st-75th | 678 (23.8%) | 124 (25.3%) | 802 (24.0%) |  |
| 76th-100th | 659 (23.1%) | 99 (20.2%) | 758 (22.7%) |  |
| **APRDRG, Likelihood of dying** |  |  |  | **<0.001\*\*\*** |
| Minor | 1293 (44.9%) | 141 (27.9%) | 1434 (42.4%) |  |
| Moderate | 858 (29.8%) | 151 (29.9%) | 1009 (29.8%) |  |
| Major | 540 (18.8%) | 158 (31.3%) | 698 (20.6%) |  |
| Extreme | 188 (6.5%) | 55 (10.9%) | 243 (7.2%) |  |
| **APRDRG, Loss of function** |  |  |  | **<0.001\*\*\*** |
| Minor (includes cases with no comorbidity or complications) | 521 (18.1%) | 57 (11.3%) | 578 (17.1%) |  |
| Moderate | 1334 (46.4%) | 197 (39.2%) | 1531 (45.3%) |  |
| Major | 843 (29.3%) | 182 (36.2%) | 1025 (30.3%) |  |
| Extreme | 180 (6.3%) | 67 (13.3%) | 247 (7.3%) |  |
| **Hospital bed size** |  |  |  | **0.012\*** |
| Small | 229 (8.0%) | 59 (11.7%) | 288 (8.5%) |  |
| Medium | 684 (23.8%) | 125 (24.9%) | 809 (23.9%) |  |
| Large | 1965 (68.3%) | 319 (63.4%) | 2284 (67.6%) |  |
| **Hospital teaching status** |  |  |  | **0.036\*** |
| Metropolitan, non-teaching | 455 (15.8%) | 103 (20.4%) | 558 (16.5%) |  |
| Metropolitan, teaching | 2308 (80.2%) | 382 (75.8%) | 2690 (79.6%) |  |
| Non-metropolitan | 114 (4.0%) | 19 (3.8%) | 133 (3.9%) |  |
| **Discharge location** |  |  |  | **<0.001\*\*\*** |
| Home self care | 2012 (69.9%) | 301 (59.8%) | 2313 (68.4%) |  |
| Home health care | 30 (1.0%) | 6 (1.2%) | 36 (1.1%) |  |
| Transfer to short-term hospital | 406 (14.1%) | 112 (22.3%) | 518 (15.3%) |  |
| Transfer to other care facility | 398 (13.8%) | 70 (13.9%) | 468 (13.8%) |  |
| Against medical advice | 32 (1.1%) | 14 (2.8%) | 46 (1.4%) |  |
| **Control/Ownership of hospital** |  |  |  | 0.850 |
| Government | 311 (10.8%) | 54 (10.7%) | 365 (10.8%) |  |
| Private non-profit | 2257 (78.4%) | 399 (79.3%) | 2656 (78.6%) |  |
| Private for-profit | 310 (10.8%) | 50 (9.9%) | 360 (10.6%) |  |
| **Hospital Designation** |  |  |  | 0.946 |
| Large metropolitan, ≥1 million residents | 1826 (63.4%) | 322 (64.0%) | 2148 (63.5%) |  |
| Small metropolitan, ≤1 million residents | 938 (32.6%) | 163 (32.4%) | 1101 (32.6%) |  |
| Micropolitan | 83 (2.9%) | 14 (2.8%) | 97 (2.9%) |  |
| Non-urban residual | 31 (1.1%) | - | 35 (1.0%) |  |

### Statistical Analysis

Statistical analysis was performed using SPSS-1.0.0.118 (IBM Corp., Armonk, N.Y., USA). Two-sided *t*-test was used for statistical analysis. Baseline characteristics between non-readmission and readmission patients were tested for statistical differences using the Pearson Chi Square test for categorical variables and Mann-Whitney U-Test for continuous variables, with non-readmission as the reference group. Multivariable logistic regression analysis was performed on all cause readmission to determine the predictors of readmission, with adjustments for age-group, gender, insurance status, hospital teaching status, and comorbidities (Table 3). Statistical significance was defined by p-value (0.05 *, 0.01 **, 0.001 ***).

## Results

### All-Cause Readmission Rate for Cerebral Cavernous Malformations (CCMs)

There were a total of 1881 admissions (3382 weighted) in the 2017 NRD database due to CCMs. 286 (504 weighted) of these patients had an unplanned readmission within 30 days of discharge. NRD recommends using weighted figures to estimate national prevalence in order to increase external validity of the analysis and protect patient privacy. The 30 day all cause readmission rate was 14.9% (13.7-16.2%) (Fig. 1). The frequency of readmission decreased over the 30 days after discharge, with most readmissions occurring within the first week (Fig. 2).

**Figure 1:**
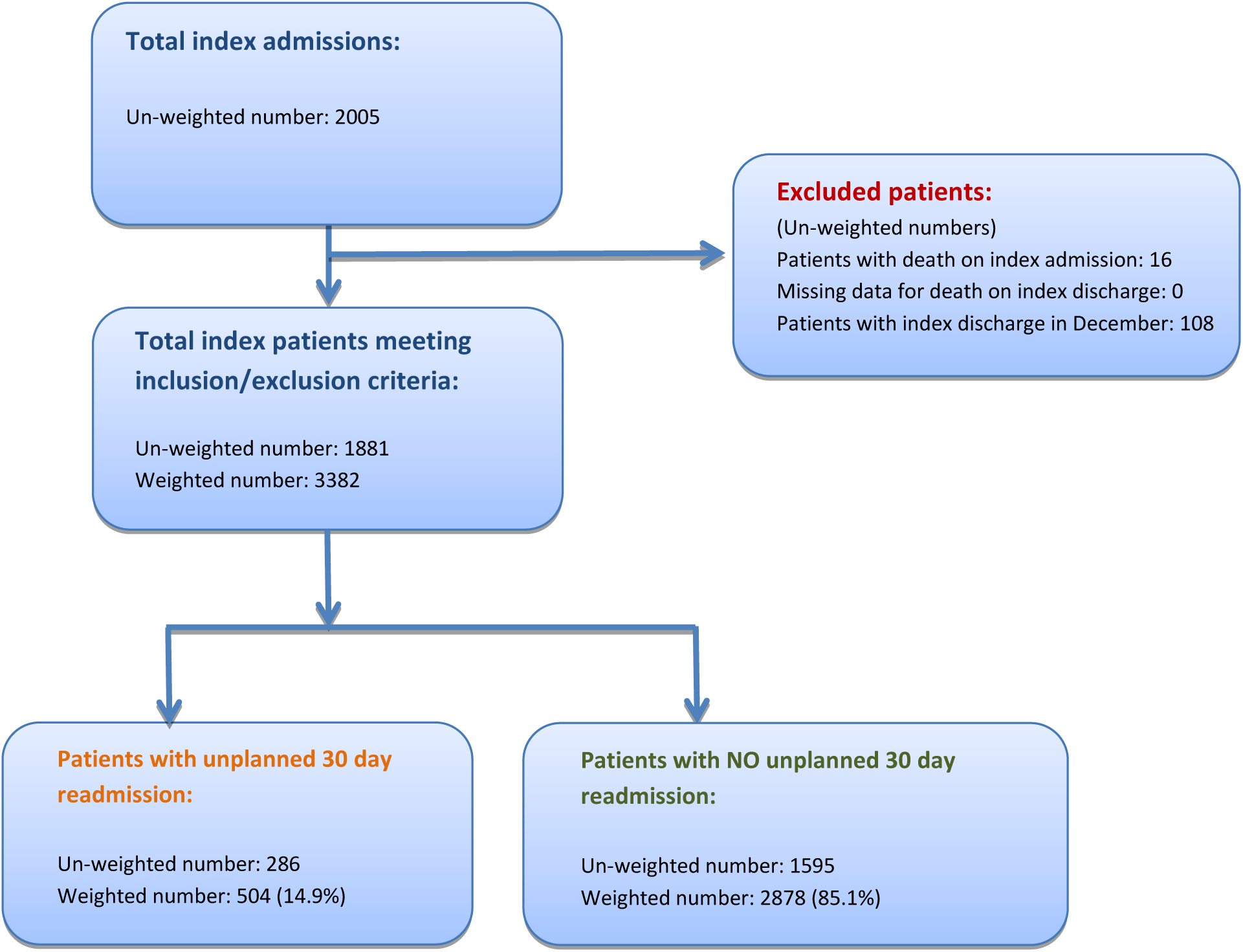
Flowchart of CCM patient selection and overall study design. Inclusion criteria are all patients admitted in US hospitals with primary diagnosis of CCM in 2017. Exclusion criteria are patients with death during the index hospitalization, missing data on index discharge, and admitted in December 2017. Patients were weighted to estimate national admission levels as recommended by Health Care Utilization Project (HCUP). Readmission is defined as any patient readmitted to the hospital within 30 days of discharge. Any patients admitted under observation and seen in the emergency room were not included.

**Figure 2:**
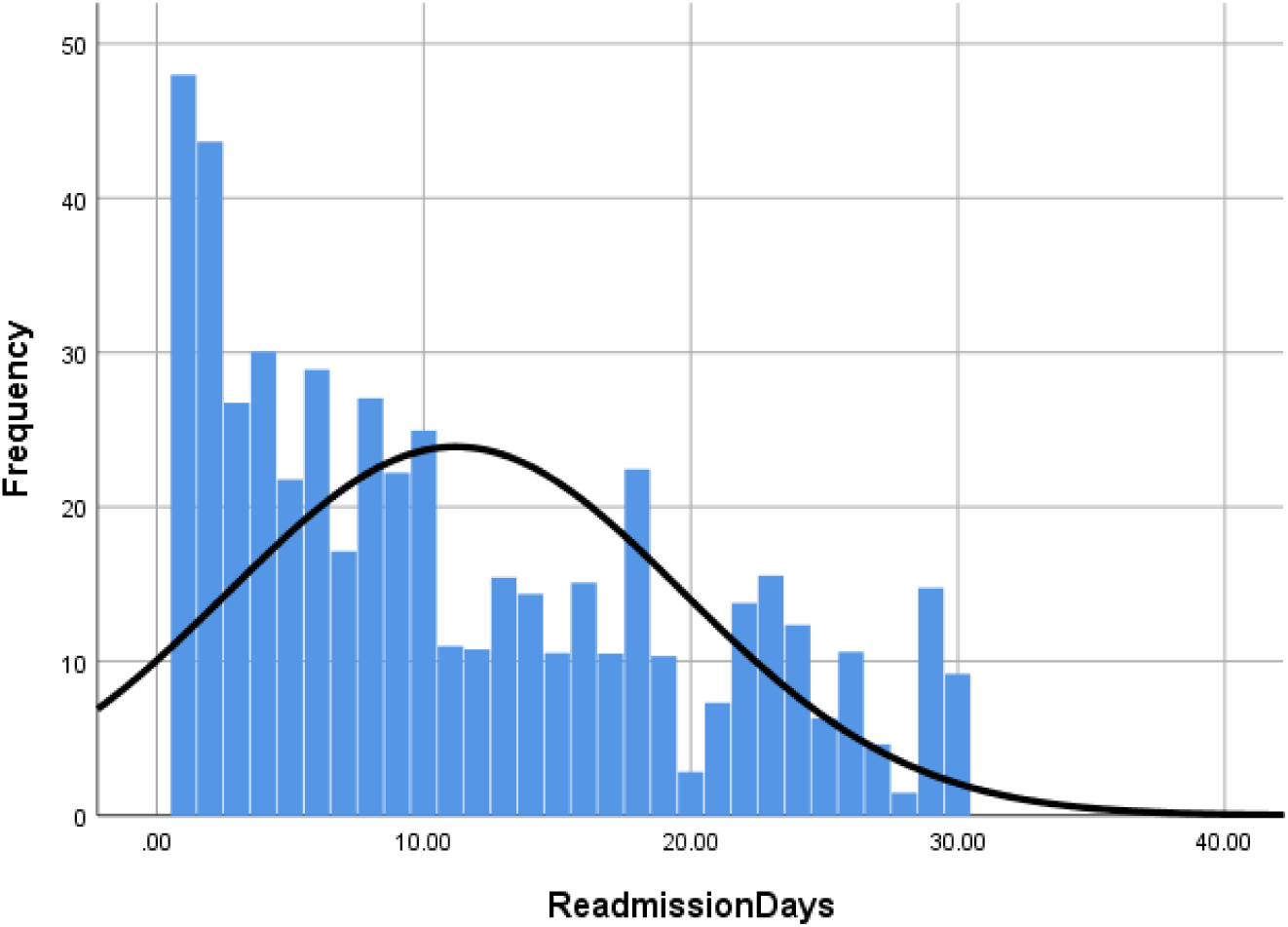
Probability Distribution Plot of All Cause Readmissions. The bar graph represents the frequency of readmissions per day after discharge from index hospitalizations. The probability distribution line graph shows the probability density of readmissions. (Mean = 11.19, Std. Dev = 8.5, N =509)

### Baseline Patient Characteristics for All Cause Readmissions

The baseline characteristics for the index hospitalization is shown in Table 1. The majority of patients in this study were women (55.7%) aged 42-72, consistent with previous reported dimorphisms of CCM severity, with increased severity in females ^15, 16^. However, there were no statistically differences seen with gender (p=0.053) and age (p=0.248). The cost of the index hospitalization (p<0.001) and length of index hospitalization (p<0.001) were significantly higher in readmitted patients. Readmitted patients were more likely to have Medicare and Medicaid and less likely to have private insurance. There were no differences in the income quartile between readmitted and non-readmitted patients. Patients with higher risk of mortality (p<0.001) and loss of function (p<0.001) were more likely to get readmitted in a severity-dependent manner. Patients seen at smaller sized hospitals were more likely to get readmitted than a larger sized hospital (p=0.012). Patients seen at non-teaching metropolitan hospitals were readmitted more frequently than teaching metropolitan hospitals (p=0.036), suggesting that quality of healthcare may have impact on readmission rates. Patients that were discharged home were less likely to get readmitted while patient set up with home health, short-term, or long-term care facility were more likely to get readmitted (p<0.001).

### Patient Comorbidities for All Cause Readmission Rates

The effect of patient comorbidities on all cause readmission rates is shown in Table 2. Statistically significant higher readmission rates were seen in patients with substance use (p=0.028), lipid disorders (p=0.046), diabetes (p<0.001), GI bleeding (p<0.001), coagulation disorders (p=0.019), renal failure (p<0.001), and CAD (p<0.001). Interestingly, significantly lower readmission rates were seen in patients with tobacco use (p=0.028), dermatological lesions (p=0.029), and headaches (p=0.039). No difference was observed in other neurological comorbidities such as seizures, ischemic stroke, and hemorrhagic stroke (Table 2).

**Table 2:** Comorbidities in CCM patients with 30 day all cause readmissions. Patient baseline comorbidities without all cause 30-day readmission (first column), with all cause 30-day readmission (second column), and total patient population (third column) are shown. The p-value for the characteristic between the two groups is also shown (fourth column). Each cell shows the weighted number of patients and percentage, as recommended by HCUP. Data from any cell with N<5 is removed as per HCUP requirements. All significant p-values are bolded. Skin lesions represent all dermatological abnormalities, non-specific for CCMs. All CCSR codes used to define the comorbidities are shown in supplementary table 1. Abbreviations: CCSR-Clinical Classifications Software Refined, CAD-coronary artery disease, GI-gastrointestinal, PAD-peripheral arterial disease. *p<0.05, **p<0.01, ***p<0.001.

| Comorbidities | No 30d readmission N (%) | 30d readmission N (%) | Overall N (%) | P value |
| --- | --- | --- | --- | --- |
| Alcohol Use | 180 (6.3%) | 24 (4.8%) | 204 (6.0%) | 0.197 |
| CAD | 365 (12.7%) | 109 (21.6%) | 474 (14.0%) | **<0.001\*\*\*** |
| Coagulation disorders | 167 (5.8%) | 43 (8.5%) | 210 (6.2%) | **0.019\*\*** |
| Diabetes | 532 (18.5%) | 142 (28.2%) | 674 (19.9%) | **<0.001\*\*\*** |
| GI Bleed | 17 (0.6%) | 11 (2.2%) | 28 (0.8%) | **<0.001\*\*\*** |
| Headache | 375 (13.0%) | 49 (9.7%) | 424 (12.5%) | **0.039\*** |
| Hypertension | 1175 (40.8%) | 213 (42.3%) | 1388 (41.0%) | 0.546 |
| Hemorrhagic stroke | 508 (17.7%) | 88 (17.5%) | 596 (17.6%) | 0.917 |
| Ischemic stroke | 273 (9.5%) | 53 (10.5%) | 326 (9.6%) | 0.470 |
| Infections | 285 (9.9%) | 48 (9.5%) | 333 (9.8%) | 0.803 |
| Lipid disorders | 928 (32.2%) | 185 (36.8%) | 1113 (32.9%) | **0.046\*** |
| Obesity | 348 (12.1%) | 71 (14.1%) | 419 (12.4%) | 0.204 |
| PAD | 106 (3.7%) | 22 (4.4%) | 128 (3.8%) | 0.454 |
| Pregnancy | 72 (2.5%) | - | 76 (2.2%) | - |
| Renal Failure | 268 (9.3%) | 84 (16.7%) | 352 (10.4%) | **<0.001\*\*\*** |
| Seizures | 758 (26.3%) | 130 (25.8%) | 888 (26.3%) | 0.798 |
| Skin lesions | 173 (6.0%) | 18 (3.6%) | 191 (5.6%) | **0.029\*** |
| Substance abuse | 152 (5.3%) | 39 (7.7%) | 191 (5.6%) | **0.028\*** |
| Tobacco use | 408 (14.2%) | 53 (10.5%) | 461 (13.6%) | **0.028\*** |

### Predictors of All Cause Readmissions

In multivariable logistic regression analysis (Table 3), only one age group (65-74) had significantly fewer readmissions than the reference population (>75). Private insurance (OR 0.441, p<0.001) was protective of readmissions, relative to Medicaid, while there were no differences seen among Medicare, self-pay, no charge, and other insurances. Metropolitan teaching hospitals (OR 0.769, p=0.039) decreased odds of readmission relative to non-teaching metropolitan hospitals. Several comorbidities including substance use (OR 1.805, p=0.003), diabetes (OR 1.335, p=0.018), GI bleeding (OR 3.427, p=0.002), renal failure (OR 1.432, p=0.027), and CAD (1.429, p=0.010) significantly increased odds of readmission, while none of the neurological comorbidities (seizures, ischemic stroke, hemorrhagic stroke, and headache) affected readmission odds.

**Table 3:** Multivariable logistic regression analysis for all cause readmission due to CCMs. Multivariate logistical regression analysis with variables of age, gender, insurance status, hospital teaching status, and comorbidities is shown. The odds ratio and its 95% confidence interval is shown (column 2). The p-value for the characteristic between the two groups is also shown (third column). The reference group for age, gender, insurance status, and hospital teaching status is designated. All significant p-values are bolded. All CCSR codes used to define the comorbidities are shown in supplementary table 1. Abbreviations: CCSR-Clinical Classifications Software Refined, CAD-coronary artery disease, GI-gastrointestinal, PAD-peripheral arterial disease. *p<0.05, **p<0.01, ***p<0.001.

| Variable | OR (95% CI) | P value |
| --- | --- | --- |
| **Age** |  |  |
| ≥75 | 1 (reference) |  |
| 18-44 | 0.720 (0.472-1.097) | 0.126 |
| 45-64 | 0.704 (0.495-1.002) | 0.051 |
| 65-74 | 0.744 (0.560-0.990) | **0.042\*** |
| **Gender** |  |  |
| Male | 1 (reference) |  |
| Female | 0.842 (0.687-1.031) | 0.096 |
| **Insurance status** |  |  |
| Medicaid | 1 (reference) |  |
| Medicare | 0.816 (0.567-1.173) | 0.272 |
| Private insurance | 0.441 (0.318-0.610) | **<0.001\*\*\*** |
| Self-pay | 0.802 (0.459-1.402) | 0.440 |
| No charge | - |  |
| Other | 0.855 (0.482-1.518) | 0.593 |
| **Hospital teaching status** |  |  |
| Metropolitan, non-teaching | 1 (reference) |  |
| Metropolitan, teaching | 0.769 (0.599-0.987) | **0.039\*** |
| Non-metropolitan | 0.665 (0.383-1.153) | 0.146 |
| **Comorbidities** |  |  |
| Tobacco use | 0.723 (0.521-1.003) | 0.052 |
| Alcohol use | 0.789 (0.497-1.253) | 0.315 |
| Substance abuse | 1.805 (1.217-2.678) | **0.003\*** |
| Lipid disorders | 0.861 (0.684-1.082) | 0.199 |
| Hypertension | 1.054 (0.839-1.324) | 0.652 |
| Diabetes | 1.335 (1.050-1.697) | **0.018\*** |
| Obesity | 1.130 (0.842-1.518) | 0.415 |
| PAD | 0.914 (0.561-1.489) | 0.718 |
| GI Bleed | 3.427 (1.544-7.603) | **0.002\*\*** |
| Coagulation disorders | 1.329 (0.918-1.922) | 0.131 |
| Infections | 0.762 (0.544-1.067) | 0.113 |
| Renal Failure | 1.432 (1.042-1.970) | **0.027\*** |
| Pregnancy | 0.474 (0.171-1.309) | 0.150 |
| CAD | 1.429 (1.090-1.872) | **0.010** |
| Seizures | 1.129 (0.895-1.424) | 0.305 |
| Ischemic stroke | 1.018 (0.732-1.416) | 0.917 |
| Hemorrhagic stroke | 1.061 (0.816-1.378) | 0.660 |
| Skin lesions | 0.682 (0.409-1.136) | 0.141 |
| Headache | 0.951 (0.683-1.325) | 0.768 |

### Etiology of 30-Day Readmissions

The primary diagnosis for each readmission is grouped by organ-based system (Table 4). As expected, the most common organ system resulting in readmission is neurological complications (33.9%). The most frequent readmission diagnosis is septicemia (8.3%), while the most frequent neurological diagnosis was acute hemorrhagic cerebrovascular disease (5.2%). The 30-day CCM lesion hemorrhage rate is 0.8% (0.5-1.2%) after index hospitalization.

**Table 4:** Etiology of All Cause 30-Day Readmission in CCMs. The majority of primary diagnoses for all cause 30-day readmission after index hospitalization are shown. The primary default CCSR category is shown for each admission. The percentage of patients readmitted for each diagnosis is detailed. The diagnoses are grouped by organ system involved.

| **Readmission Diagnosis** | **Frequency (%)** |
| --- | --- |
| **Neurological Complications** | 33.9 |
| Acute hemorrhagic cerebrovascular disease | 5.2 |
| Hemangioma of intracranial structures | 5.0 |
| Epilepsy; convulsions | 4.9 |
| Cerebral infarction | 4.3 |
| Traumatic brain injury (TBI); concussion, initial encounter | 2.4 |
| Headache; including migraine | 2.4 |
| Other specified nervous system disorders | 3.0 |
| Cardiac and circulatory congenital anomalies | 1.8 |
| Sequela of hemorrhagic cerebrovascular disease | 1.4 |
| Transient cerebral ischemia | 1.3 |
| Other specified congenital malformations of brain | 1.0 |
| Postprocedural or postoperative nervous system complication | 1.0 |
| **Infectious Etiologies** | 14.6 |
| Septicemia | 8.3 |
| Skin and subcutaneous tissue infections | 2.3 |
| Urinary tract infections | 1.7 |
| Pneumonia (except that caused by tuberculosis) | 1.2 |
| Intestinal infection | 1.0 |
| **Gastrointestinal Complications** | 5.8 |
| Gastrointestinal hemorrhage | 1.8 |
| Nausea and vomiting | 1.0 |
| Intestinal obstruction and ileus | 0.9 |
| Noninfectious gastroenteritis | 0.7 |
| Esophageal disorders | 0.7 |
| Gastrointestinal cancers - stomach | 0.7 |
| **Cardiac Complications** | 5.4 |
| Heart failure | 2.1 |
| Cardiac dysrhythmias | 1.5 |
| Syncope | 1.0 |
| Acute myocardial infarction | 0.7 |
| **Pulmonary Complications** | 4.9 |
| Chronic obstructive pulmonary disease and bronchiectasis | 1.9 |
| Acute pulmonary embolism | 1.8 |
| Respiratory failure; insufficiency; arrest | 1.2 |
| **Other complications** | 35.5 |

## Discussion

Overall, 30-day all cause readmission rate for CCM patients was 14.9%, approximately one in seven CCM patients, slightly worse than the national average hospital readmission rate (13.9%) and national readmission rate due to neurological conditions (14.0%) ^17^. To date, there is no effective medication for delaying the progression of CCM lesions, likely hindering disease management, and thereby increasing hospital readmission rates in CCM patients relative to the general population ^18^.

Female gender has been associated with worse outcomes in CCM patients ^15, 16^. This similar phenomenon was seen in the analysis in which a majority of the hospitalized patients are female (55.7%) (Table 1). However, female gender was not predictive of readmissions (Table 3). This analysis, in concordance with previous studies, suggests that women are more likely to have severe CCM disease necessitating initial hospitalizations, however both genders with active CCM lesions are equally likely to require readmission. This association between CCM disease severity and female gender has been known for decades, yet it was only recently that an explanation was discovered. One study has shown that progesterone plays a crucial role in CCM protein signaling and increased exposure to progesterone and its derivatives (progestins) may lead to increased bleeding rates of CCM lesions leading to increased hospitalizations ^19, 20^. In the general population, older patients have more frequent readmissions than younger patients. However this is not seen in CCM patient population ^17^. Accounting for comorbidities, there was a small significant decrease in readmissions in the middle-aged group (65-74) relative to the reference (Table 4). Our analysis showed that CCM readmission frequency by age is bimodal likely due to multitude of CCM genetic mutations resulting in the variable age of disease onset. Therefore, younger patients may have aggressive mutations requiring frequent readmissions while older patients, due to decreased organ reserve, may require readmission even with milder CCM phenotypes. Patients with private insurance have significantly decreased readmission rates relative to all other insurance types, which may be attributed to the increased access to outpatient resources.

Patients with readmissions had a significantly larger index hospitalization cost and index hospitalization length of stay (Table 1). This suggests that these patients had more severe presentations of the disease leading to more readmissions, which was confirmed when stratifying the patients by APRDRG functionality class and mortality class. Readmitted patients had a significantly lower baseline functionality and higher mortality risk, suggesting that severe disease leads to more readmissions. This has been shown in the general population ^17^. Patients treated at metropolitan teaching hospitals with higher bed capacity had lower readmission rates, implying that the quality of healthcare affected readmission rates (Table 3). Several large teaching hospitals in the country have been designated CCM centers for excellence, including Barrow Neurological Institute, University of Chicago, University of Virginia, University of New Mexico, Mayo Clinic, etc. All of these centers of excellence are teaching hospitals, where well-trained neurosurgeons and neurologists provide specialized care for CCM patients. This analysis suggests that treatment at these larger teaching hospitals significantly decreased readmission rates.

Certain patient comorbidities could predict which CCM patient would require readmission. Some comorbidities were seen in higher frequency in readmitted patients of which coronary artery disease (CAD), diabetes, renal failure, and substance abuse were predictive of readmission in the regression analysis (Table 2, 3). CAD ^21^, diabetes ^22^, renal failure ^23^, and substance abuse of certain drugs ^24^ have been shown to generate free radicals that could result in vascular stress and damage to certain regions of the vasculature, possibly triggering the formation of new CCM lesions. This may explain why certain lesions are more predisposed to bleeding, while other lesions remain clinically silent. Interestingly, this did not extend to all comorbidities associated with vascular stress including tobacco use and lipid disorders, suggesting that more research is needed to understand why only certain factors play major roles in readmissions. Patients with GI bleeds were found to be predictive of readmission. CCM lesions have been seen in other organs such as liver, bone, skin, and lungs but they are often only clinically significant in the CNS ^25^. Patient with history of bleeding in other organ systems may be predisposed to readmission.

The role of pregnancy in CCM disease is a disputed with some reporting no increased disease activity ^26–28^, while others suggesting there is increased activity ^29^. There are not enough pregnant patients to perform statistical testing, however no effect was seen in logistical regression analysis (Table 2,3). A proposed mechanism of CCM pathogenesis is infections and alteration of the microbiome ^30, 31^. Certain infections lead to innate immunogenic responses, such as TLR4, which was reported to result in CCM lesions ^30^. This analysis did not show any effect of infection on all cause readmission rates (Table 3). Although neurological conditions were found to be a major cause for readmission, previous history of seizures and strokes were not predictive of readmission. Patients with CCM lesions can also develop cutaneous lesions and headaches. Although headaches were seen in higher frequency among readmitted patients and dermatological lesions were seen in lower frequency among readmitted patients, there was no effect in all cause readmissions after accounting for other variables (Table 3). This is likely due to diagnoses of headache and skin lesions being non-specific for CCM disease even in CCM patients.

Analysis of all cause readmission etiologies showed that the most common primary causes of readmissions were neurological (33.9%) and infectious (14.6%). Patients with CCMs have a wide range of neurological sequalae including hemorrhagic cerebrovascular accident, seizures, and headaches ^32^. In this analysis, seizures (5.2%) and hemorrhage (4.9%) were similar in frequency. The 30-day lesion bleeding rate after index hospitalization is 0.8% (0.5-1.2%). Treatment of these sequalae requires better management of the underlying CCM disease. Hospital-acquired infectious etiologies are the second most frequent cause of readmissions in CCM patients. Hospitalized patients are prone to getting subsequent infections, and CCM patients may be at higher risk due to neurological disability requiring devices such as foleys. Timely removal of unnecessary devices may decrease readmission due to infectious etiologies. Other etiologies of all cause readmission were gastrointestinal (5.8%), cardiac (5.4%), and pulmonary (4.9%), unlikely to be related directly to CCM pathology and improved patient follow up may decrease readmissions from these etiologies.

The objective of this study is to utilize the NRD to identify risk factors associated with high risk CCM patients for 30-day readmission. If these patients are identified in the primary admission, it may be possible to allocate more resources to prevent future readmissions. Decreasing hospital readmissions not only decreases healthcare burden due to CCMs, but also improves patient quality of life. Future studies may consider using artificial intelligence and machine learning to develop algorithms and plugins for patient care software that will help clinicians implement new therapeutic strategies to improve the quality of patient care and healthcare outcomes ^13^. We also hope that by identifying similar characteristics among patients with aggressive CCM disease, it would increase understanding of the pathogenesis of CCM lesions. This methodology has been used to study other neurological diseases ^33, 34^. However, there are still limitations that must be taken in account. The NRD database is a readmission database that stores patient ICD-10 diagnosis codes for each hospitalization. Any patient that was only seen in the emergency department without hospital admission would not be included in the database. There are limitations to the elements documented in the database. Certain CCM lesion characteristics such as number of lesions, size of lesion, and location of lesion are influential in the severity of CCM disease ^35^. However, the NRD does not provide any information on these factors. Although ICD-10 codes increase the accuracy of diagnosis, inaccuracies and misdiagnoses are still possible. ICD-10 diagnostic code ‘D1802’ (hemangioma of intracranial vessels) is the most specific diagnosis for CCM, however as evidenced by readmission etiologies, few patients were diagnosed with ‘Q283’ (Other malformations of cerebral vessels). Therefore, few patients with CCM may be in this other diagnostic category. Some inaccuracies are to be expected in analyzing such large databases. Additionally, CCM is a rare disease, although its prevalence is increasing due to accessibility of MRI imaging. Even in the NRD, which encompasses 60% of all inpatient admissions in the USA, there are not enough readmitted patients with certain comorbidities such as pregnancy to adequately power the statistical test to identify a difference. Lastly, this is a retrospective observational study and therefore causation cannot be inferred from the data.

## Conclusion

This study in the first and only study analyzing readmission rates in CCM patients, with such a substantial sample size. Readmission rates have gathered popularity in the last several years and have direct financial implications to healthcare facilities and in allocation of resources. Decreasing readmissions would decrease the financial burden of CCM lesions and improve overall patient quality of life. In this study, several patient characteristics and comorbidities were identified that increased the odds of readmission. Future efforts include intervening on high-risk patients for readmission and reassessment of disease burden on CCM patients.

## Data Availability

N/A

## Abbreviations

(CCM): Cerebral Cavernous Malformations
(DVA): Developmental Venous Anomaly
(NRD): National Readmission Database
(HCUP): Healthcare Cost and Utilization Project
(IRB): Institutional Review Board
(CCSR): Clinical Classifications Software Refined
(ICD-10): International Classification of Disease
(IQR): Interquartile range
(LOS): Length of Stay
(APRDRG): All Patient Refined Diagnosis Related Groups
(PAD): Peripheral Arterial Disease
(GI): Gastrointestinal
(CAD): Coronary Artery Disease

## Legends

**Supplementary Table 1: Clinical Classifications Software Refined (CCSR) codes for CCM and associated comorbidities**

All the CCSR/ICD-10 codes used in this study for each comorbidity are shown.

