## Supplementary material for "Readmissions in Patients with Cerebral Cavernous Malformations (CCMs): A National Readmission Database (NRD) Study": Suppl Materials

Running Title: Readmission Rates in CCM

* Co-first Authors
†† Co-correspondent authors

All correspondence:
Jun Zhang, Sc.D., Ph.D.
Department of Molecular and Translational Medicine (MTM)
Texas Tech University Health Science Center El Paso
5001 El Paso Drive, El Paso, El Paso, TX 79905


**Supplementary Table 1: Clinical Classifications Software Refined (CCSR) codes for CCM and associated comorbidities**

All the CCSR/ICD-10 codes used in this study for each comorbidity are shown.

Supplementary Table 1: Clinical Classifications Software Refined (CCSR) codes for CCM and associated comorbidities

| Comorbidities | CCSR/ ICD-10CM /ICD10-PCS codes |
|---|---|
| CCM | D1802 |
| Alcohol use | MBD017 |
| CAD | CIR009-CIR011 |
| Coagulation disorders | BLD006 |
| Diabetes | END002, END003 |
| GI Bleed | DIG021 |
| Headache | NVS010 |
| Hypertension | CIR007 |
| Hemorrhagic stroke | CIR021 |
| Ischemic stroke | CIR020 |
| Infections | INF001-INF011 |
| Lipid disorders | END010 |
| Obesity | END009 |
| PAD | CIR026 |
| Pregnancy | PRG002 |
| Renal Failure | GEN003 |
| Seizures | NVS009 |
| Skin lesions | SYM014 |
| Substance abuse | MBD018, MBD019, MBD020, MBD021 |
| Tobacco use | MBD024 |
